# Methods of an Exercise is Medicine-Mediated Behavioral Counseling Intervention in Patients At-Risk for Cardiovascular Disease: The Vitalis Cardio Study

**DOI:** 10.1101/2025.09.17.25335262

**Authors:** Garrett M. Steinbrink, Jenna Springer, Lauren Tetmeyer, Katherine Mellen, Korey A. Kennelty, Heather Schacht Reisinger, Britt Marcussen, Dale S. Bond, Yin Wu, Lucas J. Carr

**Author notes:** **Corresponding Author:** Lucas J. Carr.

## Abstract

**Introduction:** Cardiovascular diseases (CVDs) are the leading causes of death in the U.S. Having a healthy dietary pattern and engaging in regular physical activity (PA) reduces the risk of developing CVDs. The U.S. Preventative Services Task Force and the American College of Sports Medicine’s Exercise is Medicine (EIM) initiatives recommend patients at risk for CVDs receive behavioral counseling interventions (BCIs). However, few patients are screened for lifestyle behaviors and connected with evidence-based BCIs.

**Purpose:** To describe (1) our approaches for identifying insufficiently active patients and connecting them to evidence-based BCIs and (2) the methods of our 12-week, theory-based BCI for insufficiently active patients.

**Methods:** A novel clinical workflow was implemented in six Family Medicine clinics to screen and refer insufficiently active patients to a 12-week BCI. The BCI is theoretically grounded in the Multi-Process Action Control (M-PAC) framework and includes health education, health coaching, and self-monitoring. The implementation and preliminary efficacy of the workflow and BCI will be evaluated with the RE-AIM framework. Implementation outcomes include clinical workflow metrics and BCI acceptability and fidelity. Preliminary efficacy includes changes in psychosocial mechanisms of action, device-based and self-reported health behaviors, and health outcomes. Efficacy outcomes are assessed at baseline, post-intervention, with a subset of outcomes assessed after 12 weeks of post-intervention follow-up.

**Conclusion:** This clinical workflow and BCI will inform the future implementation of primary care-based BCIs to reduce the risk of developing CVD in insufficiently active patients.

## INTRODUCTION

Cardiovascular diseases (CVDs) are the leading causes of death in the U.S., resulting in over 200,000 excess deaths [1] and $400 billion in healthcare expenditures annually [2]. Health-promoting lifestyle behaviors, such as consuming a healthy dietary pattern and engaging in regular physical activity (PA), reduce the risk of developing CVDs [3,4]. Still, few U.S. adults meet components of the Dietary Guidelines for Americans [5] and only 25% meet the recommended levels of PA [6]. Behavioral counseling interventions (BCIs) targeting diet and PA reduce the risk of CVD events by 20% in those with existing CVD risk factors [7]. As such, the U.S. Preventive Services Task Force (USPSTF) issued a recommendation to refer patients with CVD risk factors to BCIs [7]. These recommendations also align with the American College of Sports Medicine’s Exercise is Medicine® (EIM) initiative encouraging healthcare to treat PA as a “vital sign” by screening patients for inactivity and prescribing exercise and/or referring insufficiently active patients to evidence-based BCIs [8]. Unfortunately, few healthcare providers screen patients for these lifestyle behaviors and there is a lack of evidence-based BCIs to receive patient referrals.

EIM’s Physical Activity Vital Sign (PAVS) has been recommended as a screening tool to identify insufficiently active patients within healthcare [8]. The PAVS quickly identifies ∼70% of patients not meeting aerobic PA recommendations [9]. Patients identified as insufficiently active with the PAVS also demonstrate more CVD risk factors, compared to patients meeting recommendations [10,11]. This suggests the PAVS can be leveraged to identify and refer patients insufficiently active patients who are at risk of developing CVDs.

Despite its clinical utility, only a few large healthcare systems have implemented the PAVS into their clinical workflow to identify insufficiently active patients and refer them to BCIs [12]. Dayao and colleagues recently reported the implementation and effectiveness of an EIM-mediated health coaching intervention in five primary care clinics in southern California [12]. Insufficiently active patients who completed ≥2 phone-based health coaching sessions increased their PA over time, while patients who declined reduced their PA [12]. Although differences in PA were not statistically different between patient groups, these findings suggest that connecting insufficiently active patients to a BCI may improve PA. This is consistent with broader evidence indicating that PA referral schemes contribute to small, but clinically meaningful increases in PA [13]. Given the reliance on self-report PA and the “low-intensity” nature of this previous BCI [7], designing, implementing, and evaluating a higher-intensity intervention, utilizing device-measured PA outcomes, is warranted. Additionally, as BCIs targeting both diet and PA are recommended for those with CVD risk factors [7], moving beyond PA, exclusively, and evaluating a multi-behavior BCI is needed.

Guided by the RE-AIM (Reach, Effectiveness, Adoption, Implementation, and Maintenance) framework [14], this paper describes our methods for integrating an EIM-mediated screening and referral system into Family Medicine clinics at a large university teaching hospital in the Midwest region of the U.S. Specifically, we aim to describe our methods for (1) screening patients for insufficient activity, (2) connecting insufficiently active patients to evidence-based BCIs, (3) delivering a theory-based BCI, and (4) evaluating the implementation and preliminary efficacy of a BCI. Our long-term goal is to develop a scalable model that can be implemented at similar institutions. This may improve the care of patients with CVD risk factors and reduce CVD burden.

## METHODS

### Setting

This study took place in six Family Medicine clinics at the University of Iowa Health Care system, all located within 30 miles of Iowa City, Iowa, U.S. These clinics conduct approximately 17,000-19,000 outpatient visits per month, of which ∼1,500-2,000 of these visits are for annual wellness exams. Given its geography and large catchment area, this clinic has a relatively diverse patient population [11].

### Physical Inactivity Screening Process

Our clinical workflow to identify and refer insufficiently active patients is illustrated in Figure 1. Upon arrival at their annual wellness exam, patients are screened for physical inactivity by completing the PAVS [15]. The PAVS consists of two questions and is administered on a mobile tablet during the check-in process. First, the patient is asked, “On average, how many days per week do you engage in moderate to strenuous exercise (like a brisk walk)?”. Next, the patient is asked, “On average, how many minutes do you engage in exercise at this level?”. These two values are multiplied to estimate the number of weekly minutes of moderate-to vigorous-intensity PA (MVPA). If the patient reports ≥150 minutes of MVPA per week, they are identified as meeting the current aerobic PA guidelines, and the “Adequate Exercise” diagnostic code is populated in their medical chart. If the patient reports <150 minutes of MVPA per week, the system automatically flags the patient as not meeting aerobic PA guidelines and populates an “Inadequate Exercise” diagnostic code.

**Figure 1:**
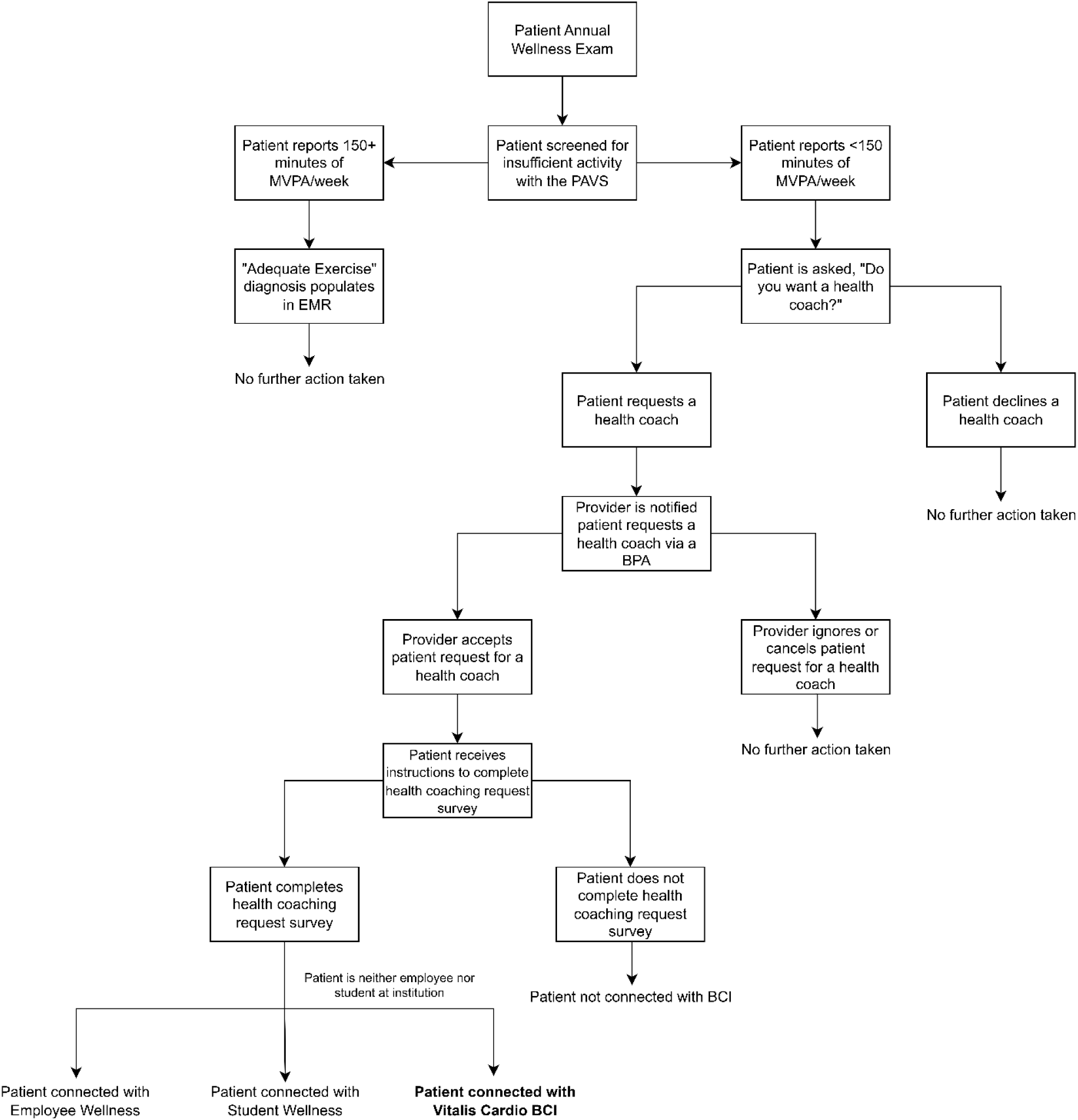
Novel clinical workflow to identify insufficiently active patients. MVPA = moderate-vigorous-intensity PA, EMR = electronic medical record, PAVS = Physical Activity Vital Sign, BPA = best-practice alert, BCI = behavioral counseling intervention

### Behavioral Counseling Intervention (BCI) Referral Process

Patients who report fewer than 150 minutes of MVPA per week are automatically asked a follow-up question, “Do you want to meet with a health coach who can help you meet your health-related goals?”. If the patient reports ‘No’, the patient proceeds with their wellness exam as normal, and no further action is taken by either the patient or provider. If the patient reported ‘Yes’, the patient’s provider receives a best practice advisory (BPA), which alerts the provider that the patient reports not meeting PA recommendations and requests to meet with a health coach. After receiving the BPA, the provider has the option to accept or cancel the BPA. If the provider accepts the BPA, a QR code to complete an online health coach request survey is added to the patient’s after visit summary (AVS) report and Epic MyChart application. If the BPA is cancelled, no health coach request survey is added to the patient’s AVS or MyChart. Patients who request a health coach and have the health coaching request survey added to their AVS and MyChart by their provider must complete a brief online (Qualtrics) health coach request survey to get connected with a local health coaching intervention. Completed health coach request surveys are automatically sent to one of three local health coach program offerings: (1) employees of the university/hospital system are connected with board-certified health coaches offered through the university’s Employee Benefits Office, (2) students of the university are connected with board-certified health coaches through the Student Wellness division at the institution, and (3) community members who are neither employees nor students at the institution are connected with a student health coach, who is enrolled in a health coaching internship program through the institution (additional details provided below).

### Student Health Coaching Training Program

Health coaching is offered to community members for free as part of a service-learning health coach training program (Table 1). The program is approved by the National Board for Health and Wellness Coaching (NBHWC) and is led by two full-time university faculty members, who are board-certified health coaches. As part of the training program, students complete two 16-week (3 credit), non-sequential courses in lifestyle medicine and principles of health coaching, followed by a 16-week (3 credit) health coaching internship. In the “Lifestyle Medicine” course, students receive an overview of evidence-based strategies to improve patient health and well-being through healthy eating, regular PA, sleep, stress management, avoidance of risky substances, and fostering positive social connections. In the “Health Coaching” course, students learn and practice evidence-based, client-centered processes (e.g., motivational interviewing) that facilitate and empower patients to develop and achieve their self-determined health and wellness goals. Students engage in weekly peer coaching practice, self-reflection, group discussions, and practice coaching sessions with clients. Students must complete both courses with a 77% or higher, complete three 30-minute coaching sessions, and receive at least 20 minutes of structured feedback from the training program director, who is a board-certified health coach. Following the completion of this coursework, students enroll in an internship where they are connected to community members interested in receiving health coaching. Students use the internship experience to achieve a portion of the 50 required health coaching sessions needed to sit for the NBHWC board certification examination.

**Table 1:** Student health coach training program.

| Training course | Pre-requisite coursework/academic progress | Recruitment and selection | Course modules and objectives | Course training methods | Training materials | Training delivery | Training evaluation | Training quality control |
| --- | --- | --- | --- | --- | --- | --- | --- | --- |
| Lifestyle Medicine (LM) | Pre-requisite coursework in (1) nutrition and health and (2) physical activity and health<br>Complete with 77% or higher in course to apply for health coaching internship | Open enrollment, capped at 35 students per section per semester; typically, junior or senior year | Motivational interviewing techniques; chronic disease; and pillars of LM (PA, nutrition, sleep, relationships, substance use, stress management) | Primarily lecture and discussion-based, hands-on coaching practice in and out of class | Readings in LM handbook; research project; 4-part coaching project; evidence-based podcasts in LM topics | In person or online (summer) | A-F grading scale; exams, participation, research project, and coaching project all factored into grade; 77% must be achieved to apply for internship | Must submit one 30-minute coaching recording to faculty and meet for at least 20 minutes of structured coaching feedback |
| Health Coaching (HC) | Pre-requisite coursework in (1) nutrition and health and (2) physical activity and health<br>Complete with 77% or higher in course to apply for health coaching internship | Open enrollment, capped at 20 students per section per semester; typically, junior or senior year | Coaching skills, coaching practice, and coaching reflection | Primarily discussion-based, minimal lecture, hands-on coaching practice in and out of class | Readings in coaching manual; 4-part coaching project; evidence-based podcasts in HC topics | In person | A-F grading scale; coaching reflections, coaching recording, discussions, participation all factored into grade; 77% must be achieved to apply for internship | Must submit one 30-minute coaching recording to faculty and meet for at least 20 minutes of structured coaching feedback |
| Health Coaching Internship | 77% or higher in both LM and HC training courses; good academic standing; and demonstrate high-quality coaching within LM pre-requisite coursework, as determined by program director | Application-based, with coaching quality as a criterion; capped at 10 students per semester | Practice coaching skills with patients; grow coaching skills in structured, supportive environment | Rigorous training program at start of semester; hands-on coaching; weekly reflection meetings with internship supervisor and health coaching interns | Primarily discussion-based; readings; videos; student-led presentations | In person | Pass/Fail grading system<br>Pass = participation, professionalism, and successful completion of course expectations | Faculty spot-checks two health coaching session recordings per semester (one at start of semester, one at mid-semester) |

### Patient Inclusion and Exclusion Criteria

Patients connected with the trained student health coach interns are eligible to receive a 12-week BCI, called *Vitalis Cardio*, if they: (1) are 18-80 years of age, (2) have at least one CVD risk factor, (3) can commit to a 12-week remotely-delivered BCI, (4) have access to a phone or computer to attend health coaching sessions, and (5) are intending on improving one or more targeted health behaviors (e.g., diet, PA). Patients are not eligible for the intervention if they: (1) cannot safely participate in PA, assessed via Physical Activity Readiness Questionnaire, (2) cannot commit to a 12-week BCI, (3) cannot access to phone or computer to complete health coaching sessions, and (4) do not intend on changing any of their health behaviors over the 12-week intervention.

### Behavioral Counseling Intervention: Theoretical Framework

This BCI was based on the Multi-Process Action Control (M-PAC) framework, a meta-theoretical framework developed to translate intentions into adopted and maintained behavior change [16]. Briefly, the M-PAC posits that action control processes (i.e., reflective, regulatory, and reflexive) are connected, progressive, and can be modified by external behavior change techniques (BCTs) to achieve long-term behavioral modification [16]. The M-PAC processes and mechanisms of action, their definitions, and specific *Vitalis Cardio* intervention components and BCTs from the taxonomy of BCTs [17] are described in Table 2.

**Table 2:** *Vitalis Cardio* BCI components and behavior change techniques (BCTs) mapped onto Multi-Process Action Control (M-PAC) framework mechanisms of action.

| M-PAC Processes | M-PAC Process Definition | Targeted M-PAC Constructs | M-PAC Construct Definitions | Behavioral counseling intervention components | Intervention BCTs<sup>17</sup><br>(Targeted M-PAC mechanisms of action) |
| --- | --- | --- | --- | --- | --- |
| **Reflective** | The conscious, deliberate expectations of performing a behavior | Instrumental attitudes | **Instrumental attitudes:**<br>evaluation of the expected benefits of behavior | Self-monitoring | (1) Provide information about behavior-health link (**instrumental attitudes**) |
|  |  | Affective attitudes | **Affective attitudes:**<br>evaluation of the expected pleasure of behavior | Health education | (2) Provide information on consequences (**instrumental attitudes, affective attitudes**) |
|  |  | Perceived capability | **Perceived capability:**<br>evaluation of one's ability to perform a behavior | Health coaching | (5) Prompt barrier identification (**perceived opportunity**) |
|  |  | Perceived opportunity | **Perceived opportunity:**<br>evaluation of the perceived social and environmental milieu that impacts behavior |  | (10) Prompt specific goal setting (**perceived opportunity, perceived capability**)<br>(13) Provide feedback on performance (**perceived capability**)<br>(23) Relapse prevention (**perceived capability**)<br>(25) Motivational interviewing (**perceived capability**) |
| **Regulatory** | The behavioral, cognitive, and affective regulation strategies used to translate intentions into behavior (i.e., action control) | Behavioral regulation | **Behavioral regulation:** one's ability to manage and control behavior | Self-monitoring | (4) Prompt intention formation (**behavioral regulation**) |
|  |  |  | **Emotional regulation:** one's ability to manage and control emotional responses to promote health-promoting behaviors | Health education | (5) Prompt barrier identification (**behavioral regulation, cognitive regulation**) |
|  |  |  | **Cognitive regulation:** one's ability to construct and regulate thoughts and beliefs to achieve intended behaviors | Health coaching | (6) Provide general encouragement (**emotional regulation**)<br>(10) Prompt specific goal setting (**behavioral regulation, cognitive regulation**) |
|  |  |  |  |  | (11) Prompt review of behavioral goals (**behavioral regulation, cognitive regulation**)<br>(12) Prompt self-monitoring of behavior (**behavioral regulation**)<br>(13) Provide feedback on performance (**behavioral regulation**)<br>(23) Relapse prevention (**behavioral regulation**)<br>(25) Motivational interviewing (**cognitive regulation, behavioral regulation**)<br>(26) Time management (**behavioral regulation**) |
| **Reflexive** | Constructs that develop from repeated action control over time | Habit | **Habit:** learned, automatic cue-behavior associations | Self-monitoring | (10) Prompt specific goal setting (**habit**) |
|  |  | Identity | **Identity:** self-categorization of oneself that influences behavior | Health coaching | (12) Prompt self-monitoring (**habit**)<br>(16) Agree on behavioral contract (**identity**)<br>(25) Motivational interviewing (**identity**) |

Our hypothesized model of health behavior and health outcomes change is illustrated in Figure 2. Briefly, we hypothesize that psychosocial mechanisms of action (e.g., perceived capability, behavioral regulation skills, habit), which are most proximal to patients, are highly sensitive to intervention. In accordance with the M-PAC framework, we contend that changes in mechanistic psychosocial outcomes lead to changes in PA, dietary, and/or sleep behaviors, which translate into improved health outcomes (e.g., blood pressure, body weight), and overall quality of life.

**Figure 2:**
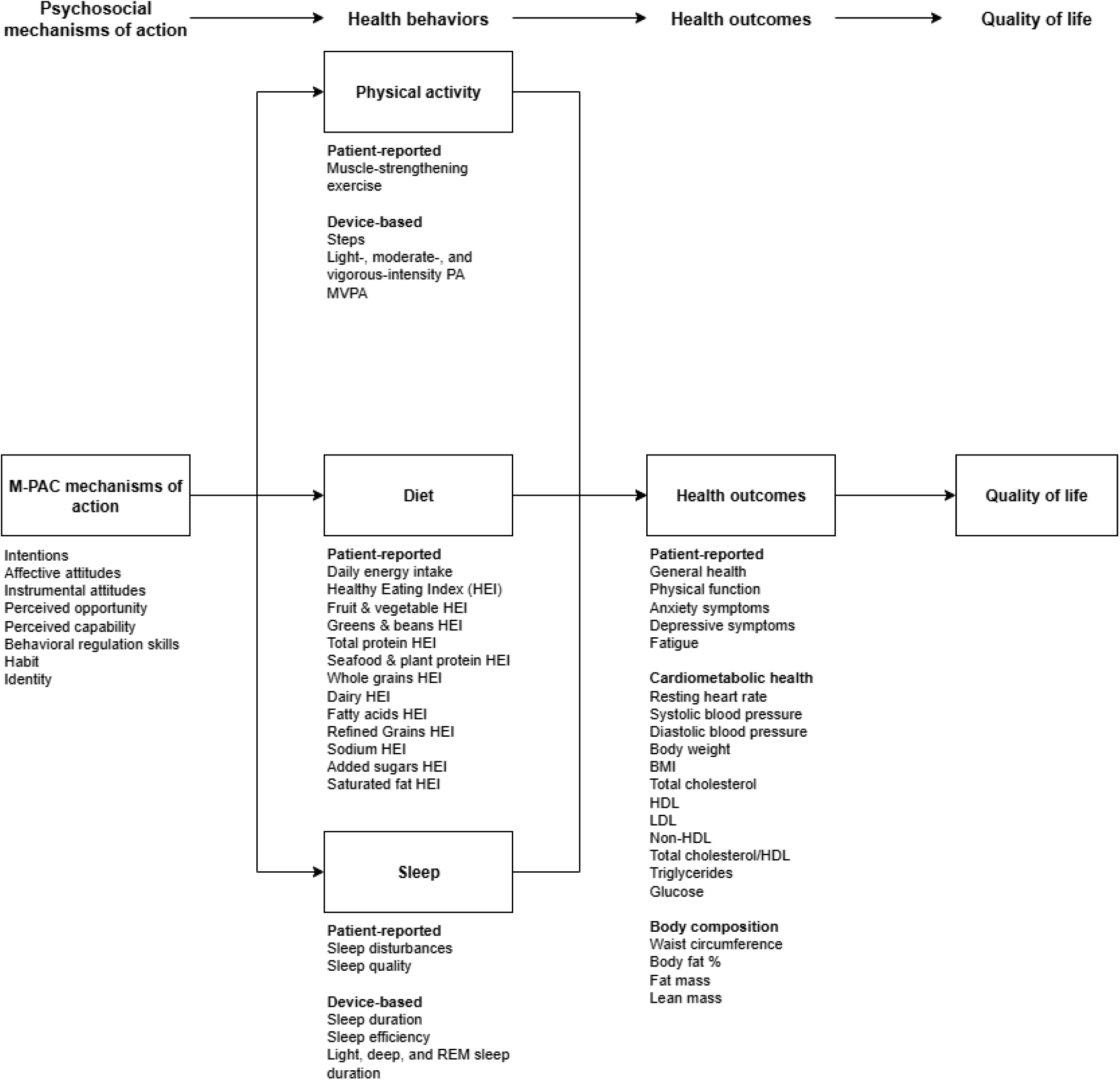
Hypothesized model of *Vitalis Cardio*.

### Behavioral Counseling Intervention: Self-Monitoring Component

Self-monitoring is a demonstrated BCT, resulting in significant improvements in diet and PA behaviors [17–19]. For this study, patients are given a wrist-worn Fitbit Inspire 3 (Google LLC, Mountain View, CA, USA), which is a commercially available PA and sleep monitoring device. Patients are also granted access to the Fitbit mobile application and encouraged to monitor their daily PA (e.g., steps, activity minutes) and sleep (e.g., sleep duration and efficiency). Patients are instructed to wear the Fitbit during the entire duration of the 12-week BCI and are also encouraged to set and monitor PA and/or sleep-related goals with their health coach. Fitbit data is uploaded and stored via the cloud-based data management service, Fitabase (Fitabase, San Diego, CA, USA). After the patient completes the 12-week intervention, they can transfer their data from the study-specific account to a personal account for future personal use.

### Behavioral Counseling Intervention: Health Education Component

Educating patients on the positive effects of health-promoting behaviors and the negative consequences of not engaging in these behaviors has been used in numerous behavior change interventions [17]. For this intervention, two Ph.D.-trained experts in exercise physiology (L.J.C) and nutrition (K.R.M) created two ∼6-8-minute videos, which educate patients about the health-promoting effects of engaging in enough PA to meet the PA Guidelines for Americans [15] and consuming a dietary pattern that is consistent with the American Heart Association’s diet and lifestyle recommendations. Patients are encouraged to watch these videos at least once during the 12-week intervention.

### Behavioral Counseling Intervention: Student Health Coaching Component

Health coaching interventions improve PA, dietary behaviors, and stress management in adults with CVD risk factors [20] and student health coaches have also been used successfully to improve these outcomes in young, college-aged adults [21]. In this intervention, patients meet with a trained student health coach remotely via a HIPAA-compliant web application (Healthie Inc., New York, NY, USA). Patients meet with their health coach 5 times, during weeks 1, 2, 4, 6, and 10 of the 12-week BCI (Figure 3). Briefly, the week 1 session lasts 90 minutes and involves the health coach: (1) prompting the patient to set a health and wellness vision for their future self, (2) assisting the patient in identifying barriers and developing strategies to remove and/or overcome them, (3) facilitating the identification of the patient’s strengths and social support, (4) helping the patient set short (i.e., weekly) and long-term (i.e., 12-week) SMART (specific, measurable, attainable, realistic, and time-based) goals, and (5) assessing the patient’s confidence/self-efficacy in achieving these short- and long-term goals. Subsequent sessions 2-4 last roughly 20 minutes and focus on: (1) identifying the patient’s psychosocial, behavioral, and/or health-related successes since the previous HC session, (2) discussing what the patient learned in pursuit of their identified goals, (3) helping the patient modify or set new weekly goals, (4) evaluating the patient’s confidence in achieving these new goals, and (5) assessing the patient’s motivation towards achieving their health and wellness vision. The fifth and final session lasts approximately 45 minutes and includes: (1) identifying what the patient learned during the intervention, (2) re-visiting the patient’s health and wellness vision and why it is important to them, (3) helping the patient set 12-week maintenance goals, and (4) assisting the patient in developing strategies to help independently achieve these maintenance goals. Importantly, because health coaching is a patient-centered process, patients are free to set any behavioral goal including, but not limited to, improving PA, diet, and/or sleep-related behaviors.

**Figure 3:**
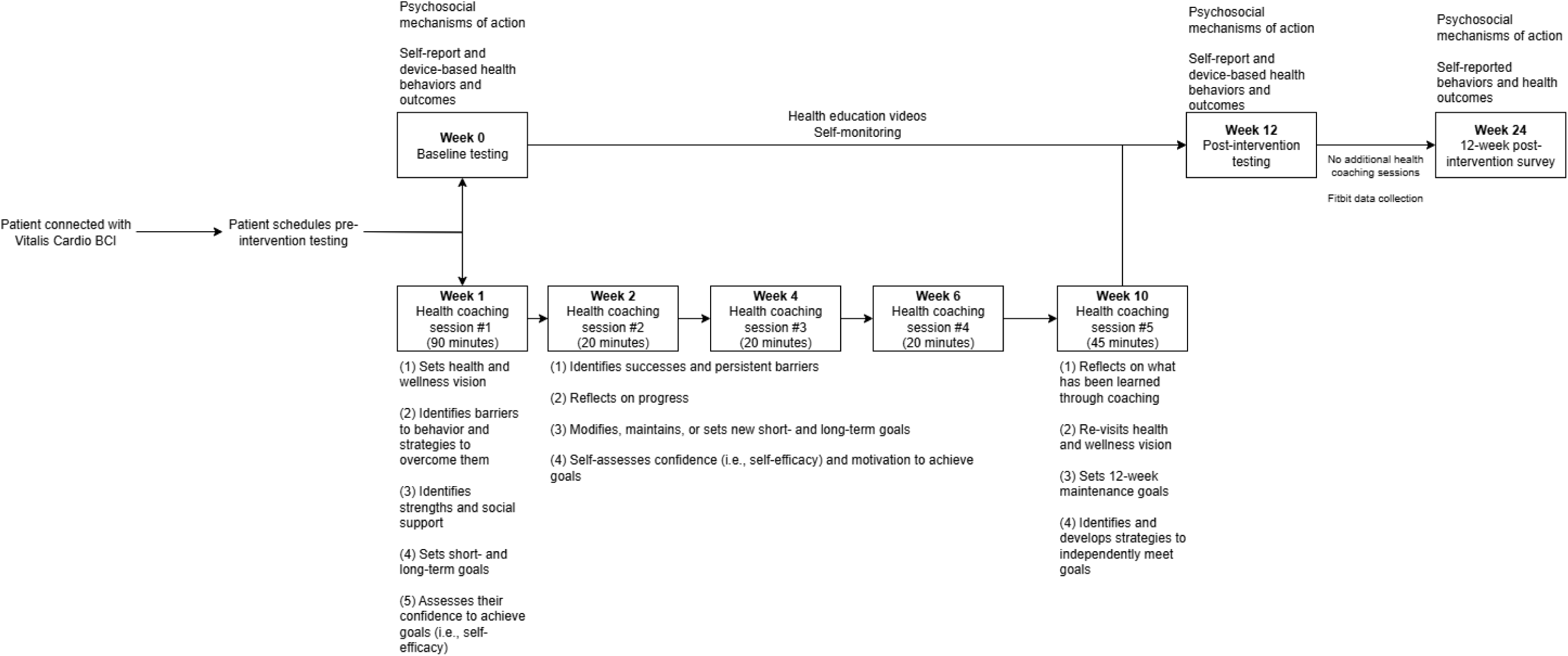
V*i*talis *Cardio* timeline.

### Behavioral Counseling Intervention: Timeline

The timeline for this BCI is illustrated in Figure 3. After patients are connected to the *Vitalis Cardio* BCI, they schedule an initial visit where they are informed of the purpose of the study and provide their informed consent with a trained research assistant. Following consent, they complete a 1–2-hour-long laboratory testing session to provide pre-intervention data for the study’s outcomes of interest. One week later, patients complete their first health coaching session. The remaining health coaching sessions are completed during weeks 2-10. On week 12, patients complete another laboratory testing session to provide post-intervention data. From weeks 13-23, no health coaching sessions are completed. Participants are, however, encouraged to wear their Fitbit during this period, to provide device-based PA and sleep follow-up data. During week 24, an online survey is sent to patients to collect a subset of self-reported outcomes, which are described below.

### Outcomes: Psychosocial mechanisms of action

Psychosocial mechanisms of action of the M-PAC framework are reported in Table 3 and were measured with 5-point Likert scales via an online survey (Qualtrics, Provo, UT, USA). Specifically, affective and instrumental attitudes, behavioral intentions, perceived capability, perceived opportunity, behavioral regulation skills, habit, and identity were measured within the context of each health behavior of interest (i.e., PA, diet, and sleep).

**Table 3:**
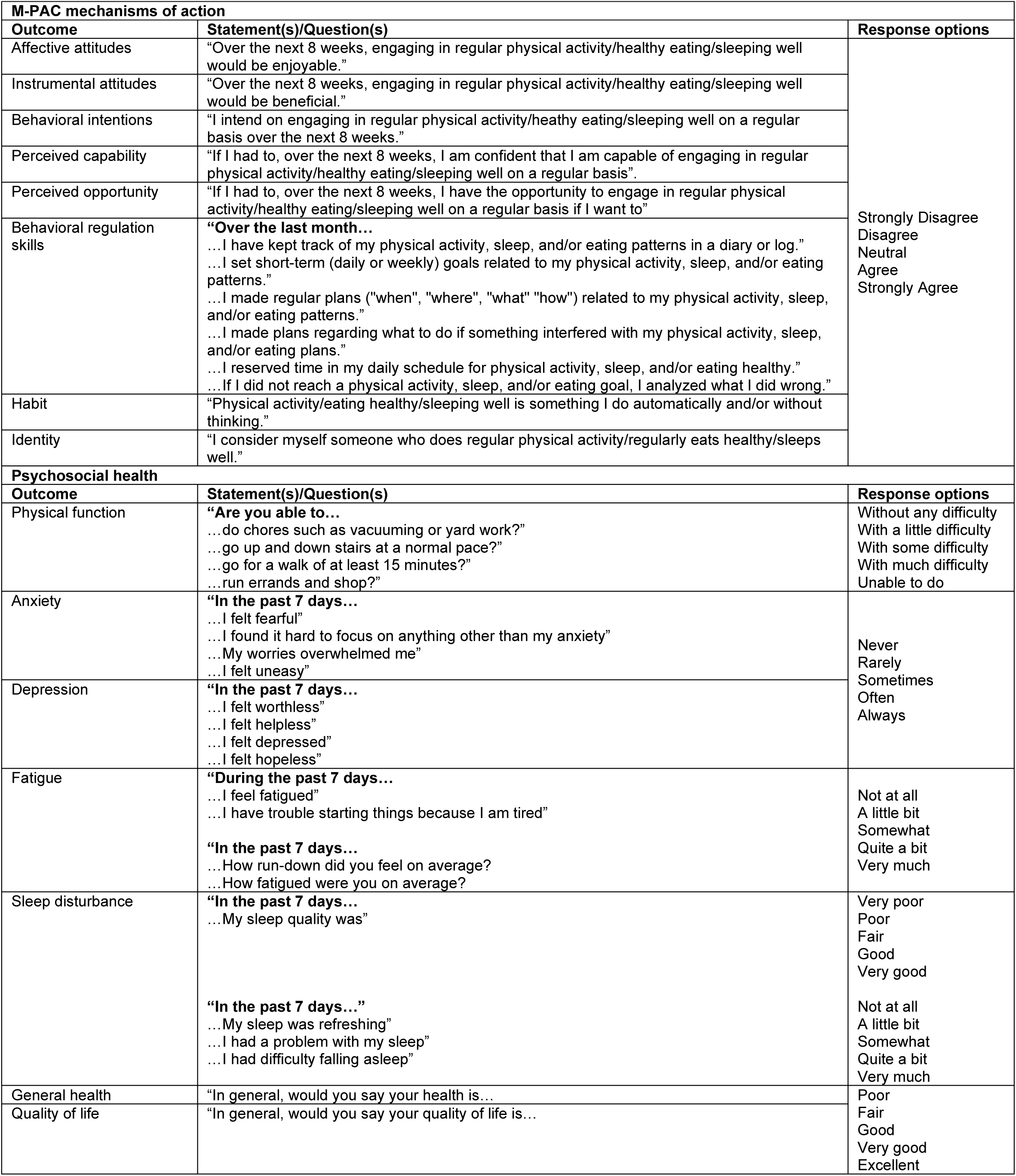
M-PAC mechanisms of action and patient-reported health outcomes of *Vitalis Cardio*.

### Outcomes: Health Behaviors

Device-based PA is measured with the Fitbit Inspire 3 at baseline, post-intervention (week 12), and after 12 weeks of post-intervention follow-up (week 24). The Fitbit demonstrates strong relationships with tri-axial accelerometry in assessing free-living MVPA [22]. Using Fitbit’s proprietary algorithms, daily number of steps and daily minutes of light-, moderate-, and vigorous-intensity PA are estimated.

Muscle-strengthening exercise (MSE) is measured with the Muscle-Strengthening Exercise Questionnaire (MSEQ) short form, which is validated against a 7-day MSE diary [23]. From the MSEQ, the weekly number of days of MSE are estimated. Adherence to current MSE guidelines is defined as engaging in MSE on at least 2 days/week [15].

Patient dietary intake is assessed with the National Cancer Institute’s (NCI) Automated Self-Administered 24-hour Recall (ASA24) at baseline and post-intervention. The ASA24 is a feasible self-report measure of previous-day dietary intake [24], and shows a high level of agreement to interview-administered 24-hour recall [25]. ASA24 data are analyzed using the *dietaryindex* R package, which demonstrates near-perfect agreement with code published by the NCI [26]. Primary dietary outcomes include estimated daily kilocalories (kcal), and overall and domain-specific (e.g., fruit, vegetables, saturated fat) Health Eating Index-2020 (HEI-2020) scores.

Device-based sleep metrics are also assessed with the Fitbit Inspire 3. Specifically, total sleep time, sleep efficiency, and time spent in specific sleep stages (i.e., light, deep, and REM sleep) are estimated with Fitbit’s proprietary sleep algorithms. The Fitbit Inspire 3 showcases good-excellent reliability in estimating total sleep duration, compared to polysomnography [27]. The single-item Sleep Quality Scale is used to assess self-report sleep quality by asking patients, “During the past 7 days, how would you rate your sleep quality overall?”, with a 10-point Likert’s scale ranging from terrible (0), to excellent (10) [28]. Sleep disturbance is assessed with the Patient-Reported Outcomes Measurement Information System-29 (PROMIS-29), which is described in Table 3.

### Outcomes: Health Outcomes

Resting systolic and diastolic blood pressure (BP) are measured using an automated OMRON HEM-907XL blood pressure monitor (OMRON Healthcare, Inc., Hoffman Estates, IL, USA) placed on the patient’s left arm. Patients sit quietly with their arm at chest level and feet flat on the floor for at least five minutes prior to BP measurement. Immediately after BP assessment, resting heart rate (HR) is measured with a HealthTree JKS50B pulse oximeter (The Health Tree Limited, Cobham, England). Waist circumference (WC) is measured at the navel using a Gulick measuring tape three times to the nearest 0.5 cm. The average of the three measurements is used for analysis. Body composition is assessed via bioelectrical impedance analysis using the Tanita MC 780U multi-frequency segmental body composition analyzer (TANITA Corporation of America, Issaquah, WA, USA). Specifically, total body mass, fat mass, lean mass in kilograms, and body fat as a percentage are estimated from the analyzer. This analyzer demonstrates high levels of agreement with dual-energy X-ray absorptiometry in assessing body fat % and fat-free mass [29]. Patients are asked to complete the test in a state of normal hydration status.

Patient metabolic profiles are measured after an overnight fast by a Cholestech LDX Analyzer (Abbott Point of Care, Princeton, NJ, USA). Total plasma cholesterol, high-density lipoprotein (HDL), total cholesterol/HDL ratio, low-density lipoprotein (LDL), non-HDL, triglycerides, and fasting glucose are assessed.

PROMIS-29 is also used to evaluate patient-reported physical function, anxiety, depression, and fatigue (Table 3). Patient-reported general health and quality of life is measured on a 5-point Likert scale.

### Outcomes: Acceptability and implementation

The acceptability and implementation outcomes are comprehensively reported in Table 4. We assess acceptability and implementation on both the patient- and the clinical workflow level. Outcomes are measured with a variety of data sources including pre- and post-intervention surveys, cloud-based data storage platforms (e.g., Fitabase), and EMR data.

**Table 4:**
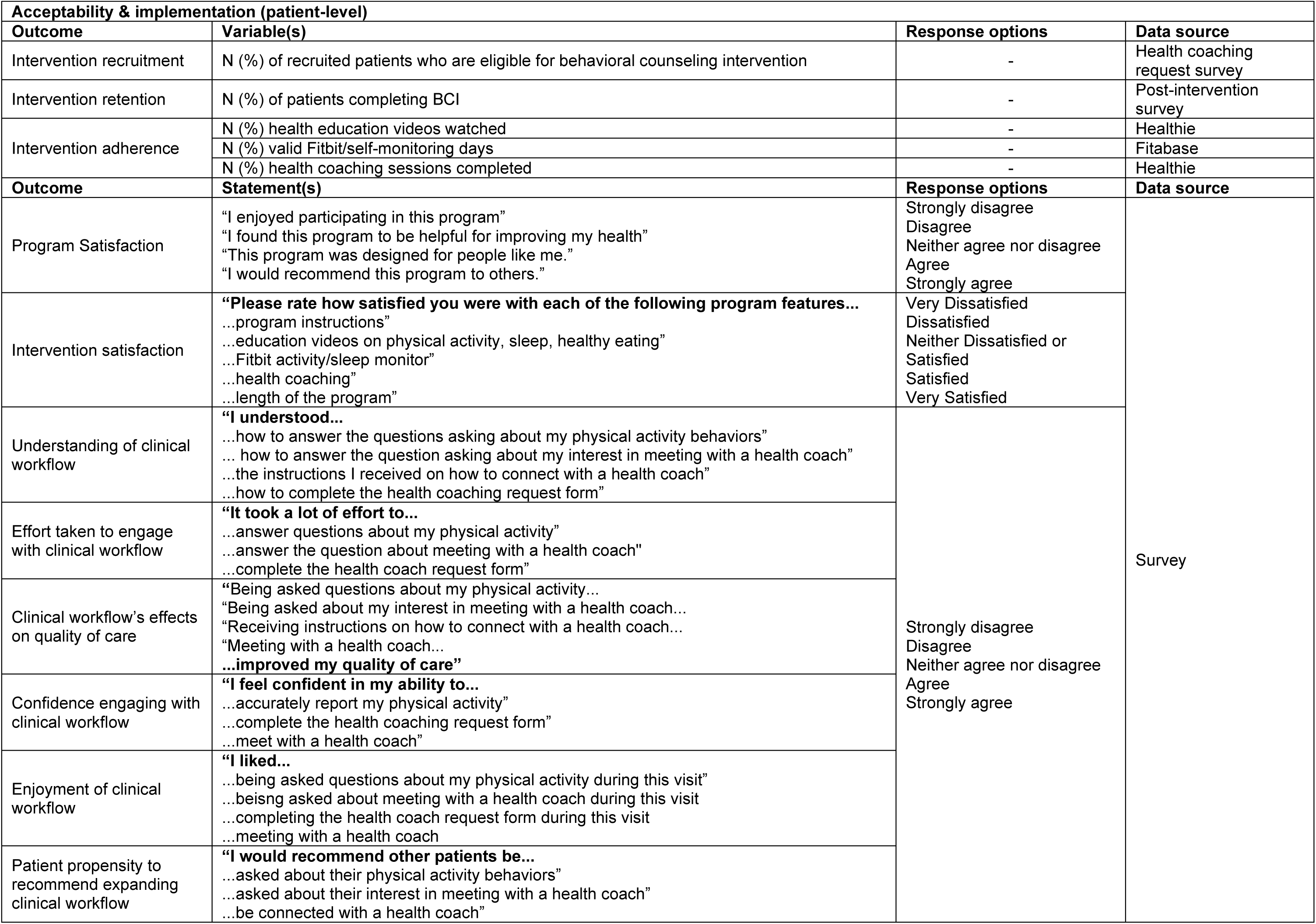
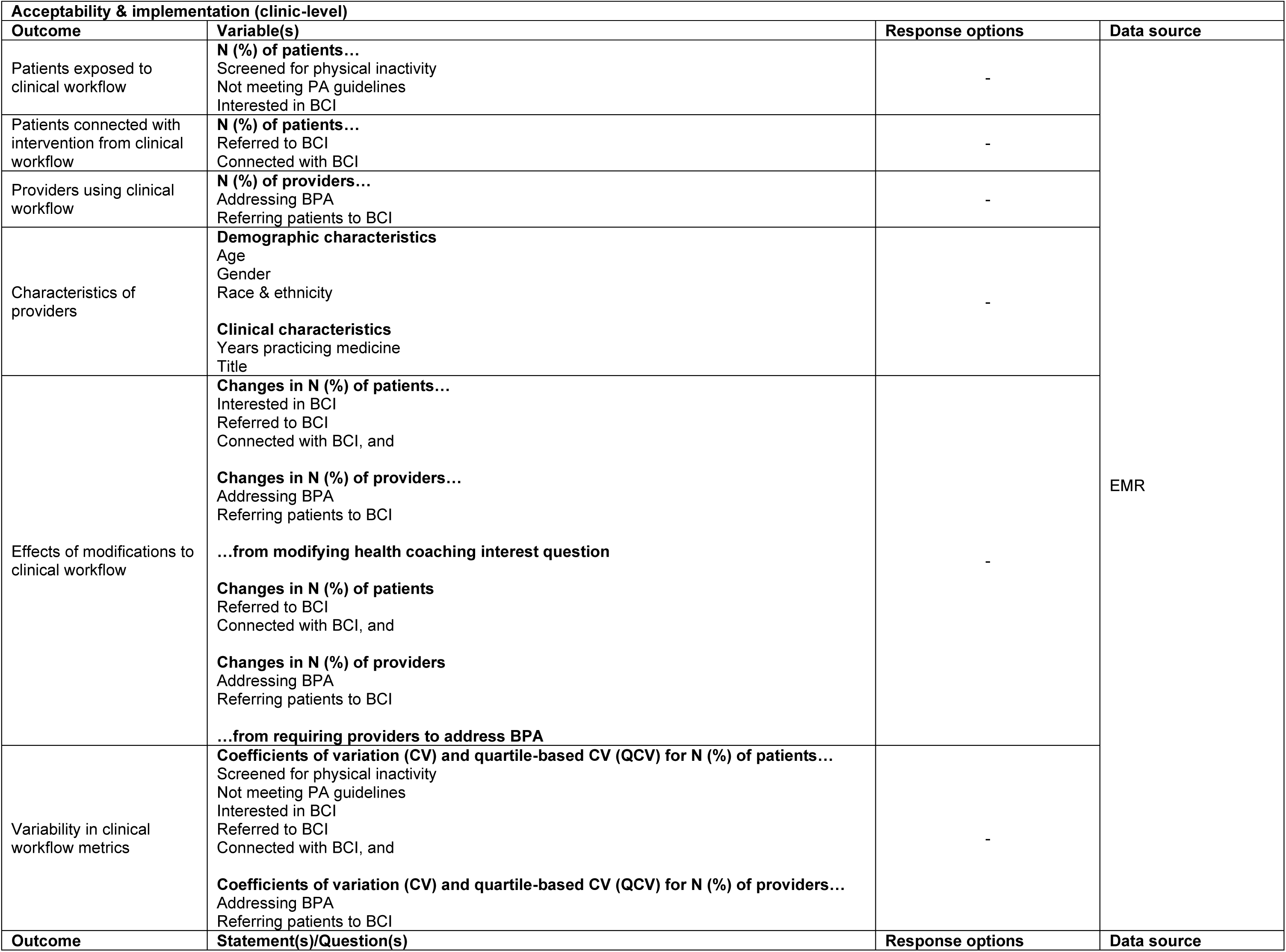
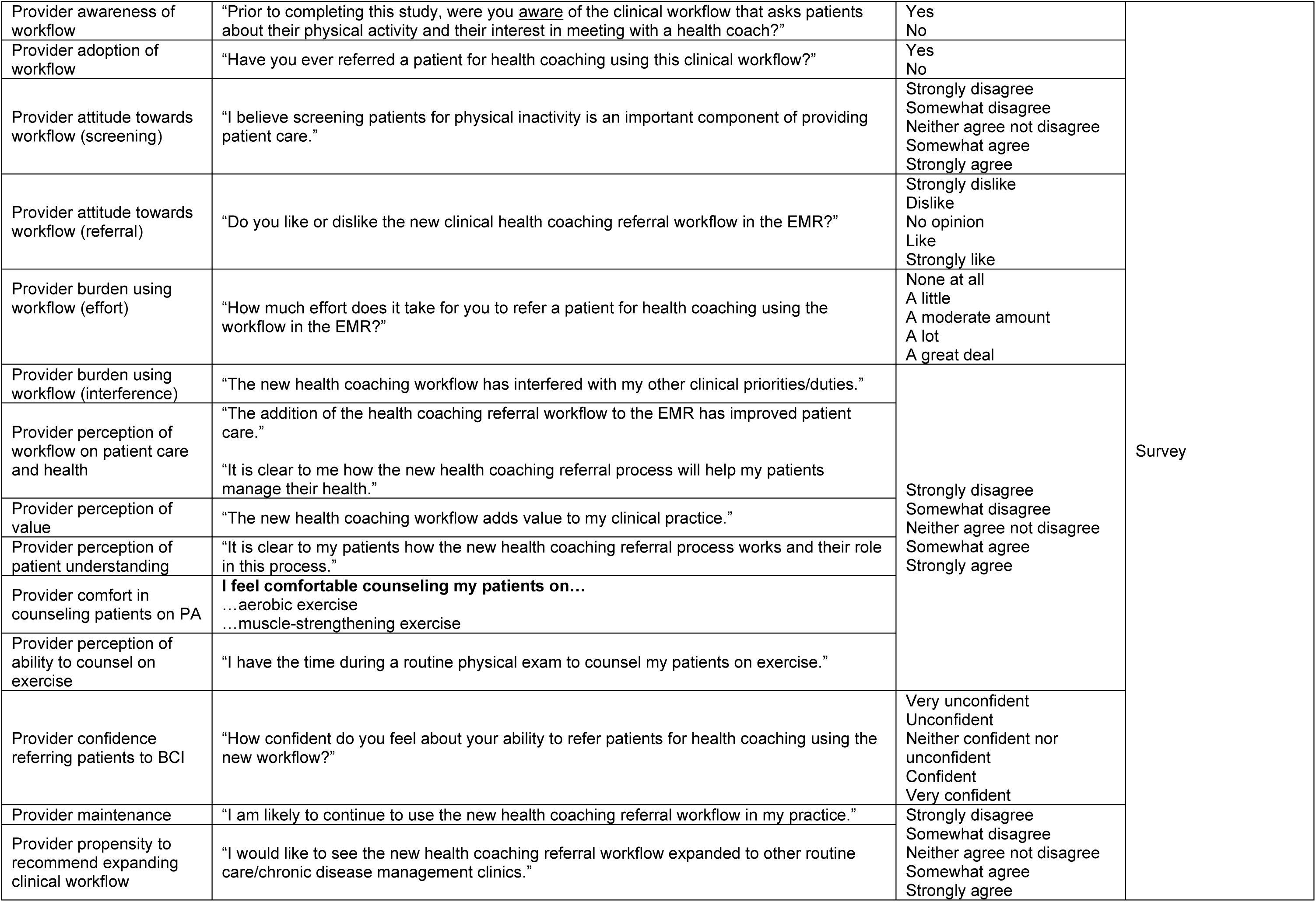
Acceptability and implementation outcomes of *Vitalis Cardio*.

### Statistical analysis

Based on our pilot work and previous studies using health coaching as a main BCI component to change PA [30], we estimated an increase in patients’ daily steps by a mean (SD) of 1,400 (2,300) from pre-post intervention. Therefore, with a power = 0.8 and an alpha = 0.05, 102 patients were needed to detect a statistically significant change of this magnitude. With an expected 20% attrition rate, we sought to recruit 128 patients to enroll in this intervention. Descriptive statistics will be used to assess the impact of and changes to the clinical workflow. For all psychosocial mechanisms of action, behavioral outcomes, and health outcomes, paired t-tests and the Chi-square test, with adjustments for multiple comparisons, will be used to assess changes from baseline to post-intervention for continuous and categorical outcomes, respectively. General linear models will be used to assess changes from baseline, post-intervention (week 12), and after 12 weeks of post-intervention follow-up (week 24). Given we will not be adequately powered to appropriately evaluate changes in all measured outcomes, effect sizes will also be calculated and reported.

### RE-AIM evaluation framework

We will use the RE-AIM framework to guide the evaluation of our clinical workflow and intervention. A comprehensive overview of our outcomes within the RE-AIM framework is shown in Table 5. Briefly, ‘Reach’ will consist of describing how many patients were exposed to our EIM clinical workflow, as well as the characteristics of patients who enrolled and completed the BCI. The effectiveness of our clinical workflow to accurately identify and refer insufficiently active patients interested in a BCI will be assessed. The short- and long-term preliminary efficacy of the intervention to elicit changes in psychosocial mechanisms of action, health behaviors, and health outcomes will also be evaluated. ‘Adoption’ will be evaluated by assessing how many providers refer patients to the BCI using this clinical workflow. To assess ‘Implementation’, we will evaluate the acceptability and fidelity of the intervention. Additionally, we will assess the effects of modifications on the clinical workflow on RE-AIM dimensions. Finally, we will evaluate both patient and provider acceptability to the clinical workflow and solicit recommendations for future refinement of the workflow and BCI. ‘Maintenance’ will be assessed by describing the variability in clinical workflow outcomes, and by patient-reported psychosocial mechanisms of action, behaviors, and health outcomes after 12 weeks of post-intervention follow-up.

**Table 5:** All *Vitalis Cardio* outcomes, guided by the RE-AIM framework.

| RE-AIM Dimension | Outcomes | Evaluation time point |  |  |  |
| --- | --- | --- | --- | --- | --- |
|  |  | Continuously | Baseline | Post-Intervention | 12-Week Post-Intervention |
| Reach | **Clinical workflow** |  |  |  |  |
|  | N (%) patients screened for physical inactivity in clinics | X |  |  |  |
|  | N (%) patients not meeting PA guidelines | X |  |  |  |
|  | N (%) patients interested in *Vitalis Cardio* | X |  |  |  |
|  | **Demographic characteristics** |  |  |  |  |
|  | Age |  | X |  |  |
|  | Gender identity |  | X |  |  |
|  | Education |  | X |  |  |
|  | Race & ethnicity |  | X |  |  |
|  | Household income |  | X |  |  |
|  | Marital status |  | X |  |  |
| Effectiveness | **Clinical workflow** |  |  |  |  |
|  | N (%) patients referred to BCI | X |  |  |  |
|  | N (%) patients connected with *Vitalis Cardio* |  | X |  |  |
|  | **Psychosocial mechanisms of action** |  |  |  |  |
|  | Behavioral intentions |  | X | X | X |
|  | Attitudes (instrumental, affective) |  | X | X | X |
|  | Perceived capability |  | X | X | X |
|  | Perceived opportunity |  | X | X | X |
|  | Behavioral regulation skills |  | X | X | X |
|  | Habit |  | X | X | X |
|  | Identity |  | X | X | X |
|  | **Behaviors** |  |  |  |  |
|  | Self-report sleep quality |  | X | X | X |
|  | Self-report sleep disturbance |  | X | X | X |
|  | Self-report MSE |  | X | X | X |
|  | ASA24 caloric intake |  | X | X | X |
|  | ASA24 HEI-2020 scores |  | X | X | X |
|  | Fitbit sleep duration |  | X | X | X |
|  | Fitbit sleep efficiency |  | X | X | X |
|  | Fitbit light sleep |  | X | X | X |
|  | Fitbit deep sleep |  | X | X | X |
|  | Fitbit REM sleep |  | X | X | X |
|  | Fitbit steps |  | X | X | X |
|  | Fitbit LPA |  | X | X | X |
|  | Fitbit MPA |  | X | X | X |
|  | Fitbit VPA |  | X | X | X |
|  | Fitbit MVPA |  | X | X | X |
|  | **Health** |  |  |  |  |
|  | Self-report physical function |  | X | X | X |
|  | Self-report anxiety symptoms |  | X | X | X |
|  | Self-report depression symptoms |  | X | X | X |
|  | Self-report fatigue |  | X | X | X |
|  | Self-report general health |  | X | X | X |
|  | Self-report QoL |  | X | X | X |
|  | Resting SBP |  | X | X |  |
|  | Resting DBP |  | X | X |  |
|  | Resting HR |  | X | X |  |
|  | Height |  | X | X |  |
|  | Weight |  | X | X |  |
|  | BMI |  | X | X |  |
|  | Bodyfat % |  | X | X |  |
|  | Total fat mass |  | X | X |  |
|  | Total lean mass |  | X | X |  |
|  | Waist circumference |  | X | X |  |
|  | Total cholesterol |  | X | X |  |
|  | Total HDL |  | X | X |  |
|  | Total triglycerides |  | X | X |  |
|  | Total LDL |  | X | X |  |
|  | Total non-HDL |  | X | X |  |
|  | Total LDL/HDL |  | X | X |  |
|  | Glucose |  | X | X |  |
| Adoption | **Clinical workflow** |  |  |  |  |
|  | N (%) providers addressing BPA | X |  |  |  |
|  | N (%) providers referring patients to BCI | X |  |  |  |
| Implementation | **Clinical workflow** |  |  |  |  |
|  | Modifications in clinical workflow on changes in RE-AIM outcomes | X |  |  |  |
|  | Provider and patient acceptability of clinical workflow | X |  |  |  |
|  | **Intervention recruitment & retention** |  |  |  |  |
|  | N (%) of recruited patients who are eligible for *Vitalis Cardio* |  | X |  |  |
|  | N (%) of enrolled participants completing *Vitalis Cardio* |  |  | X |  |
|  | **Intervention adherence** |  |  |  |  |
|  | N (%) of health education videos watched |  |  | X |  |
|  | N (%) of valid self-monitoring days |  |  | X |  |
|  | N (%) of health coaching sessions completed |  |  | X |  |
| Maintenance | **Clinical workflow** |  |  |  |  |
|  | Variability in N (%) patients screened for physical inactivity | X |  |  |  |
|  | Variability in N (%) patients not meeting PA guidelines | X |  |  |  |
|  | Variability in N (%) patients interested in BCI | X |  |  |  |
|  | Variability in N (%) patients referred to BCI | X |  |  |  |
|  | Variability in N (%) patients connected with *Vitalis Cardio* | X |  |  |  |
|  | Variability in N (%) providers addressing BPA | X |  |  |  |
|  | Variability in N (%) providers referring patients to BCI | X |  |  |  |
|  | **Psychosocial mechanisms of action** |  |  |  |  |
|  | Behavioral intentions |  |  |  | X |
|  | Instrumental attitudes |  |  |  | X |
|  | Affective attitudes |  |  |  | X |
|  | Perceived capability |  |  |  | X |
|  | Perceived opportunity |  |  |  | X |
|  | Behavioral regulation skills |  |  |  | X |
|  | Habit |  |  |  | X |
|  | Identity |  |  |  | X |
|  | **Behaviors** |  |  |  |  |
|  | Self-report sleep quality |  |  |  | X |
|  | Self-report sleep disturbance |  |  |  | X |
|  | Self-report MSE |  |  |  | X |
|  | ASA24 caloric intake |  |  |  | X |
|  | ASA24 HEI-2020 scores |  |  |  | X |
|  | Fitbit sleep duration |  |  |  | X |
|  | Fitbit sleep efficiency |  |  |  | X |
|  | Fitbit light sleep |  |  |  | X |
|  | Fitbit deep sleep |  |  |  | X |
|  | Fitbit REM sleep |  |  |  | X |
|  | Fitbit steps |  |  |  | X |
|  | Fitbit LPA |  |  |  | X |
|  | Fitbit MPA |  |  |  | X |
|  | Fitbit VPA |  |  |  | X |
|  | Fitbit MVPA |  |  |  | X |
|  | **Health** |  |  |  |  |
|  | Self-report physical function |  |  |  | X |
|  | Self-report anxiety symptoms |  |  |  | X |
|  | Self-report depression symptoms |  |  |  | X |
|  | Self-report fatigue |  |  |  | X |
|  | Self-report general health |  |  |  | X |
|  | Self-report QoL |  |  |  | X |

PA = physical activity, BCI = behavioral counseling intervention, MSE = muscle-strengthening exercise, REM = rapid eye movement, LPA = light-intensity physical activity, MPA = moderate-intensity physical activity, VPA = vigorous-intensity physical activity, MVPA = moderate-vigorous-intensity physical activity, SBP = systolic blood pressure, DBP = diastolic blood pressure, HR = heart rate, BMI = body mass index, HDL = high-density lipoprotein, LDL = low-density lipoprotein, BPA = best-practice advisory, RE-AIM = Reach, Effectiveness, Adoption, Implementation, Maintenance, QoL = quality of life

## DISCUSSION

The purpose of this methods paper was to describe our process for implementing and evaluating a BCI for insufficiently active patients at risk for CVD delivered through primary care. Specifically, we described our process for identifying and referring insufficiently active patients to a robust, evidence-based, and theoretically grounded BCI, which incorporates health education, health coaching, and self-monitoring components. Moreover, we described the theoretical framework for the intervention and mapped both intervention components and BCTs onto this framework. Finally, using the validated RE-AIM evaluation framework, we reported the psychosocial, behavioral, health outcomes evaluated at baseline, post-intervention, and selected outcomes after 12 weeks of post-intervention follow-up, to determine the preliminary efficacy of this intervention. The acceptability and implementation outcomes of this BCI are also measured and will be reported in forthcoming publications. The over-arching findings from this project will inform the development and implementation of evidence-based BCIs in the primary care setting, to improve care and reduce the individual and population-level burdens of CVD.

## Data Availability

There was no data produced or reported for this manuscript. Future data produced from the study will be available upon reasonable request to the authors.

## Informed consent

Informed consent was obtained from all individual participants included in the study.

## Ethical approval

All procedures performed are in accordance with the ethical standards of the institutional and/or national research committee and with the 1964 Helsinki declaration and its later amendments or comparable ethical standards.

## Clinical trial registration

This study is registered on ClinicalTrials.gov (NCT07001774).

